# Cesarean section prevalence at a baby-friendly hospital in southern Brazil: current context in the face of COVID-19

**DOI:** 10.1101/2021.05.03.21256516

**Authors:** Manoela de Azevedo Bicho, Mayra Pacheco Fernandes, Luís Paulo Vidaletti, Juliana dos Santos Vaz

## Abstract

**Objective:** To analyze cesarean prevalence at a baby-friendly hospital in Southern Brazil between 2017 and 2020 and possible annual and monthly changes due to the novel coronavirus (COVID-19) pandemic.

**Methods:** Descriptive cross-sectional study using secondary data retrieved from the electronic information system of a Baby-Friendly Hospital in the municipality of Rio Grande, RS, Brazil. Data was retrieved for all hospitalizations at the obstetric center between January 1^st^ 2017 and December 31^st^ 2020. Data on COVID-19 deaths were obtained from the municipal government website. Annual and monthly cesarean prevalence rates were calculated in comparison to the same periods in 2017, 2018 and 2019. Differences in prevalence rates were tested using a chi-square test, taking a significance level of less than 5%. Prevalence ratios were estimated for 2018/2017, 2019/2018, and 2020/2019.

**Results:** 7,294 childbirths were included. Cesarean prevalence was 42.6% in 2017, 44.3% in 2018, 40.2% in 2019, and 51.0% in 2020. In 2018/2017, there was no statistically significant difference in cesarean prevalence (95%CI: 0.90-1.20). Between 2019/2018, there was a reduction of approximately 20% in prevalence (95%CI: 0.69-0.93). The scenario changes between 2020/2019 with a 40% increase in cesareans (95%CI: 1.20-1.62). The period comprising July to December 2020 was the only period in which over half the deliveries were done by cesarean section, exceeding 60% in July.

**Conclusion:** Cesarean prevalence rates increased in 2020 in relation to the three preceding years. The data highlight the need to reinforce compliance with childbirth protocols to reduce cesarean sections in baby-friendly hospitals.

## Introduction

Concern has been growing in the last 30 years about the increased prevalence of cesarean section and its consequences for maternal and child health^1^. In Brazil, cesarean prevalence rose from 15% in the 1970s^2^ to 55% in 2016, accounting for more than half the number of childbirths^3^. Studies have suggested that cesarean sections have been used in an indiscriminate manner, without considering the medical reasons that justify them^7,9^. Cesarean section is a recognized barrier to early initiation of breastfeeding, and it is also associated with negative consequences in the short and long term, in particular maternal mortality, postpartum infection, neonatal mortality, newborn respiratory problems, prematurity, low weight at birth, risk of being hospitalized in an intensive care unit and ^8,10-12^.

The Baby-Friendly Hospital Initiative (BFHI) is a global initiative to promote, protect and support breastfeeding. The implementation of the “Ten Steps to Successful Breastfeeding” requires cesarean surgery rates less than or equal to 30% or development of a plan to reduce cesarean rates to 10% per year^4-6^. Thus, in a baby-friendly hospital, cesarean section must be indicated in specific situations when there is an obstetric risk associated to chilbirth^7,8^.

In March 2020, the pandemic caused by the coronavirus (COVID-19) depleted health systems globally, imposing changes in work routines in hospitals, including neonatal and maternity units^13,14^. Protocols previously adopted in maternity hospitals to promote care at birth and neonatal care underwent modifications to reduce the time spent by mothers and babies in hospital with the aim of minimizing the likelihood of exposure to the virus^15,16^.

Early in the pandemic, COVID-19 status itself became a common indication for cesarean delivery^17^. A systematic review of 203 peer-reviewed case studies of pregnant women who tested positive for SARS-CoV-2 before delivery found that 68.9% were submitted to cesareans despite lack of evidence of mother-to-child transmission. That study highlighted the need to uphold current recommendations from trusted organizations as new data are published to prevent the increase of unnecessary and unplanned cesarean sections during the pandemic^17^.

The adjustments of protocols for childbirth due to the COVID-19 pandemic along with the growing epidemic of cesareans can promote a serious increase in cesarean rates worldwide, especially in Brazil. Studies on this issue have not been identified so far in Brazil. This study aimed to analyze cesarean prevalence at a baby-friendly hospital in Southern Brazil between 2017 and 2020, identifying possible annual and monthly changes due to the COVID-19 pandemic.

## Methods

### Design, target population and sample

This is a descriptive cross-sectional study using data from the electronic information system of Dr. Miguel Riet Corrêa Jr., University Hospital of the Universidade Federal do Rio Grande (HU-FURG). This hospital is in the municipality of Rio Grande in the state of Rio Grande do Sul, Brazil. The municipality has around 211,005 inhabitants (2019) and a municipal human development index of 0.744 (available at: https://www.ibge.gov.br/cidades-e-estados/rs/rio-grande.html). In 2019, a total of 2,753 childbirths was registered in the municipality, of which 65% occurred at HU-FURG.

HU-FURG is a public institution and is part of the Brazilian Hospital Service Company, it only provides services to Brazilian National Health System Unified. In 2002, the hospital was accredited as a Baby-Friendly Hospital, offering to mothers and babies all support actions needed for maternal breastfeeding recommended by the BFHI. Currently, it is the only hospital in the extreme south of Rio Grande do Sul to be accredited as a Baby-Friendly Hospital. In the pandemic, this hospital became the reference in COVID treatment in the municipality.

For the present study, we selected all pregnant women who gave live birth at the HU-FURG maternity hospital. The exclusion criteria were those women registered as HIV-positive, cases of miscarriages or stillbirth, and birth of twins.

### Data source

Complete information was retrieved on all hospitalizations in the obstetrics center registered on the HU-FURG electronic system for the period January 1st, 2017 to December 31^st^ 2020. The data used refer to information on child delivery and characteristics of live born babies and their respective mothers.

Information on COVID-19 deaths between January 31^st^ and December 31^st^ 2020 in the municipality of Rio Grande was retrieved from the daily newssheets available on the city government website (available at: https://www.riogrande.rs.gov.br/corona/)^18^. The information contained in the newssheets is provided by the epidemiological bulletins published by the State Health Departments. The data are updated daily when the bulletins are published at 11 p.m. Brasilia time.

### Operationalization of the variables

The dependent variable was cesarean section (no; yes), according to the mode of delivery. The following variables were considered to describe the sample: maternal age (≤ 19, 20-29, ≥30 years), sex of the newborn (female; male), gestational age assessed according to the Capurro method (<37, 37-38, 39-41 and ≥42 weeks), weight at birth (<2500, 2500-2999, 3000-3500, 3501-4000 and >4000 grams) and birth shift hours [(day: from 7 a.m. to 6.59 p.m.) or at night (from 7 p.m. to 6.59 a.m.)]. The year of birth variable (2017, 2018, 2019 and 2020) was created based on date of birth.

### Data analysis

To compare cesarean prevalence in 2020 with the preceding years, we extracted the data from January 1^st^ to December 31^st^ for the years 2017, 2018 and 2019. The data on hospitalizations at the obstetrics center held on the HU-FURG electronic system were exported to Excel® and statistical analyses were performed using Stata statistical package version 16.0.

We performed descriptive analyses of the independent and dependent variables to describe the sample. Cesarean prevalence rates were estimated for each year and its respective months. Differences between annual and monthly prevalence rates were tested using the chi-square heterogeneity test, taking a significance level of less than 5%. We also calculated the prevalence ratios for cesarean deliveries between the years 2018 and 2017; 2019 and 2018; 2020 and 2019. Crude and adjusted prevalence ratios (PR) and their respective confidence intervals were calculated using Poisson regression with robust adjustment for variance. Adjustments were performed to eliminate the effect of confounding factors. All variables were taken to the multivariate model, and those with a value of ≤ 0.20 were maintained.

### Ethical aspects

The research project was submitted and approved by the Research Ethics Committee of the Universidade Federal de Pelotas (Approved file N° 4.281.378) and by the Section of Teaching and Research Administration of the HU-FURG. Database was obtained with deidentified data, and analyses were conducted in accordance with the principles of the Declaration of Helsinki.

## Results

A total of 7,744 births occurred between 2017 and 2020. After applying the exclusion criteria, 7,294 births were included. The demographic characteristics of the sample and mode of delivery are shown in Table 1. Fifty-two per cent of women were 20-29 years, and 68% of deliveries occurred between the 39^th^ and 41^st^ gestational weeks. Thirty-eight per cent of the newborn babies weighed between 3000 and 3500 grams and 58% of deliveries were performed during the day.

**Table 1.**
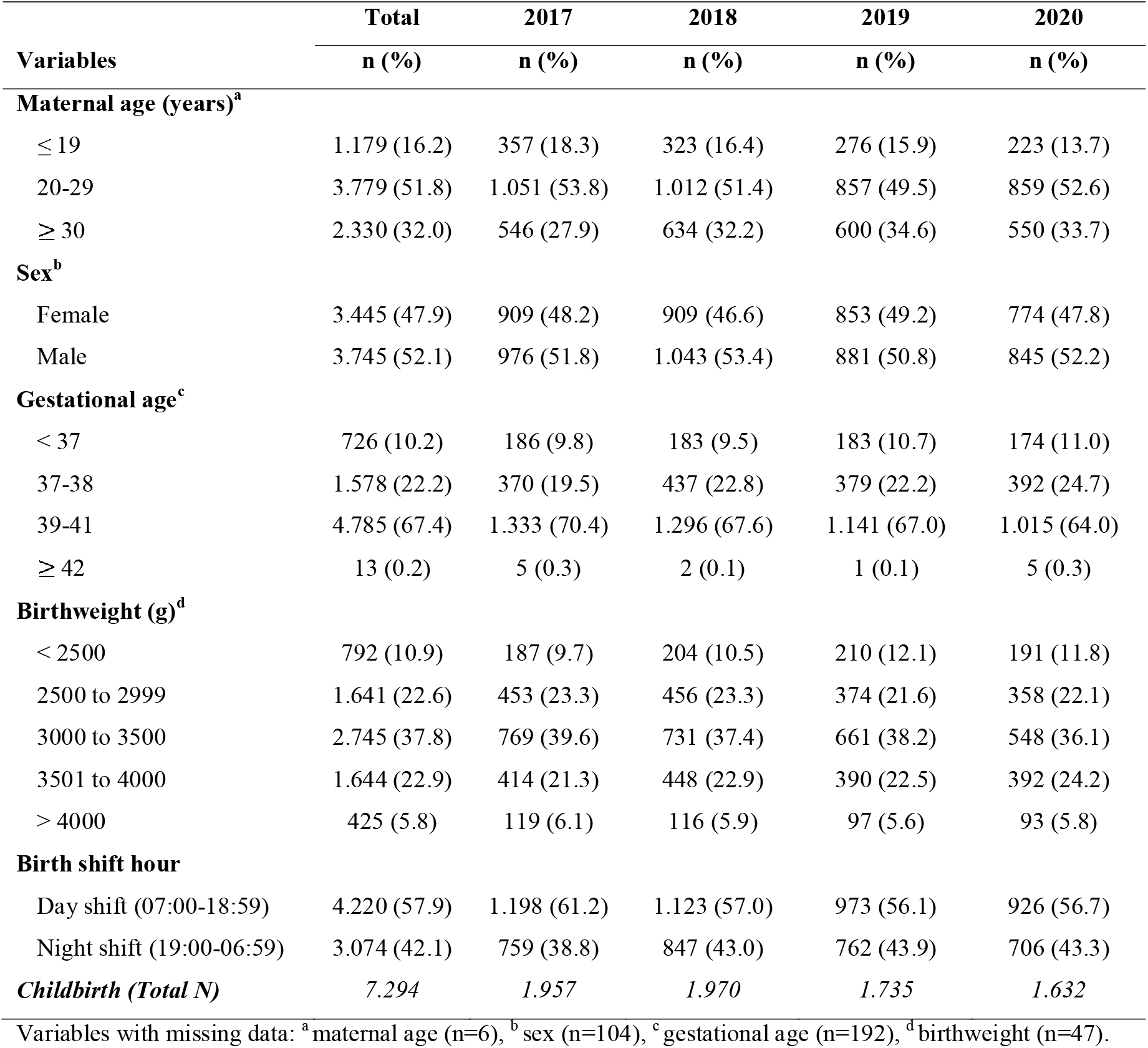
Maternal, newborn and childbirth characteristics in a baby-friendly hospital. Rio Grande, RS, Brazil (2017-2020).

| **Variables** | **Total** | **2017** | **2018** | **2019** | **2020** |
| --- | --- | --- | --- | --- | --- |
|  | **n (%)** | **n (%)** | **n (%)** | **n (%)** | **n (%)** |
| **Maternal age (years)<sup>a</sup>** |  |  |  |  |  |
| ≤ 19 | 1.179 (16.2) | 357 (18.3) | 323 (16.4) | 276 (15.9) | 223 (13.7) |
| 20-29 | 3.779 (51.8) | 1.051 (53.8) | 1.012 (51.4) | 857 (49.5) | 859 (52.6) |
| ≥ 30 | 2.330 (32.0) | 546 (27.9) | 634 (32.2) | 600 (34.6) | 550 (33.7) |
| **Sex<sup>b</sup>** |  |  |  |  |  |
| Female | 3.445 (47.9) | 909 (48.2) | 909 (46.6) | 853 (49.2) | 774 (47.8) |
| Male | 3.745 (52.1) | 976 (51.8) | 1.043 (53.4) | 881 (50.8) | 845 (52.2) |
| **Gestational age<sup>c</sup>** |  |  |  |  |  |
| < 37 | 726 (10.2) | 186 (9.8) | 183 (9.5) | 183 (10.7) | 174 (11.0) |
| 37-38 | 1.578 (22.2) | 370 (19.5) | 437 (22.8) | 379 (22.2) | 392 (24.7) |
| 39-41 | 4.785 (67.4) | 1.333 (70.4) | 1.296 (67.6) | 1.141 (67.0) | 1.015 (64.0) |
| ≥ 42 | 13 (0.2) | 5 (0.3) | 2 (0.1) | 1 (0.1) | 5 (0.3) |
| **Birthweight (g)<sup>d</sup>** |  |  |  |  |  |
| < 2500 | 792 (10.9) | 187 (9.7) | 204 (10.5) | 210 (12.1) | 191 (11.8) |
| 2500 to 2999 | 1.641 (22.6) | 453 (23.3) | 456 (23.3) | 374 (21.6) | 358 (22.1) |
| 3000 to 3500 | 2.745 (37.8) | 769 (39.6) | 731 (37.4) | 661 (38.2) | 548 (36.1) |
| 3501 to 4000 | 1.644 (22.9) | 414 (21.3) | 448 (22.9) | 390 (22.5) | 392 (24.2) |
| > 4000 | 425 (5.8) | 119 (6.1) | 116 (5.9) | 97 (5.6) | 93 (5.8) |
| **Birth shift hour** |  |  |  |  |  |
| Day shift (07:00-18:59) | 4.220 (57.9) | 1.198 (61.2) | 1.123 (57.0) | 973 (56.1) | 926 (56.7) |
| Night shift (19:00-06:59) | 3.074 (42.1) | 759 (38.8) | 847 (43.0) | 762 (43.9) | 706 (43.3) |
| **Childbirth (Total N)** | **7.294** | **1.957** | **1.970** | **1.735** | **1.632** |

Variables with missing data: <sup>a</sup> maternal age (n=6), <sup>b</sup> sex (n=104), <sup>c</sup> gestational age (n=192), <sup>d</sup> birthweight (n=47).

Regarding the annual prevalence of cesarean, rate was 42.6% in 2017, 44.3% in 2018, 40.2% in 2019, and 51.0% in 2020 (p<0.001). July to December 2020 was the only period over four years in which more than half the deliveries were cesarean, exceeding 60% in July (Table 2).

**Table 2.** Prevalence of cesarean section in a baby-friendly hospital. Rio Grande, RS, Brazil (2017-2020). ^a^

| Cesarean/month | 2017 | 2018 | 2019 | 2020 | p-value<sup>b</sup> |
| --- | --- | --- | --- | --- | --- |
|  | n (%) | n (%) | n (%) | n (%) |  |
| January | 57 (41.0) | 67 (36.8) | 57 (39.9) | 72 (49.0) | 0.157 |
| February | 51 (32.9) | 57 (33.1) | 45 (37.5) | 69 (46.4) | **0.039** |
| March | 77 (40.3) | 65 (43.6) | 61 (36.3) | 74 (52.9) | **0.027** |
| April | 78 (48.2) | 77 (49.4) | 50 (36.0) | 70 (42.4) | 0.081 |
| May | 77 (49.7) | 80 (43.5) | 55 (36.7) | 60 (44.8) | 0.149 |
| June | 67 (41.6) | 87 (51.8) | 53 (37.9) | 73 (47.7) | 0.154 |
| July | 75 (42.1) | 101 (53.2) | 69 (42.6) | 78 (60.0) | 0.065 |
| August | 87 (45.3) | 70 (43.2) | 66 (42.0) | 69 (50.0) | 0.538 |
| September | 73 (48.0) | 71 (42.3) | 53 (36.3) | 68 (56.2) | **0.009** |
| October | 54 (40.3) | 66 (46.5) | 73 (43.2) | 72 (57.6) | **0.029** |
| November | 67 (40.6) | 62 (44.0) | 60 (54.0) | 64 (54.2) | 0.054 |
| December | 70 (40.5) | 70 (44.9) | 55 (42.6) | 63 (55.3) | 0.087 |
| **Cesarean (Jan-Dec)** | **833 (42.6)** | **873 (44.3)** | **697 (40.2)** | **832 (51.0)** | **<0.001** |

<sup>a</sup> Total of cesarean deliveries, n= 4059;
<sup>b</sup> p-value refers to the chi-squared test.

Figure 1 shows the evolution of cesarean sections according to the number of COVID-19 deaths in the year 2020. Between January and December, an increase in cesarean prevalence can be seen over the months. In July there was a marked increase in the number of deaths in the municipality, accompanied by an increase in cesarean prevalence. In September and October, the cesarean curve rose, whereas the COVID-19 death rate fell sharply.

**Figure 1.**
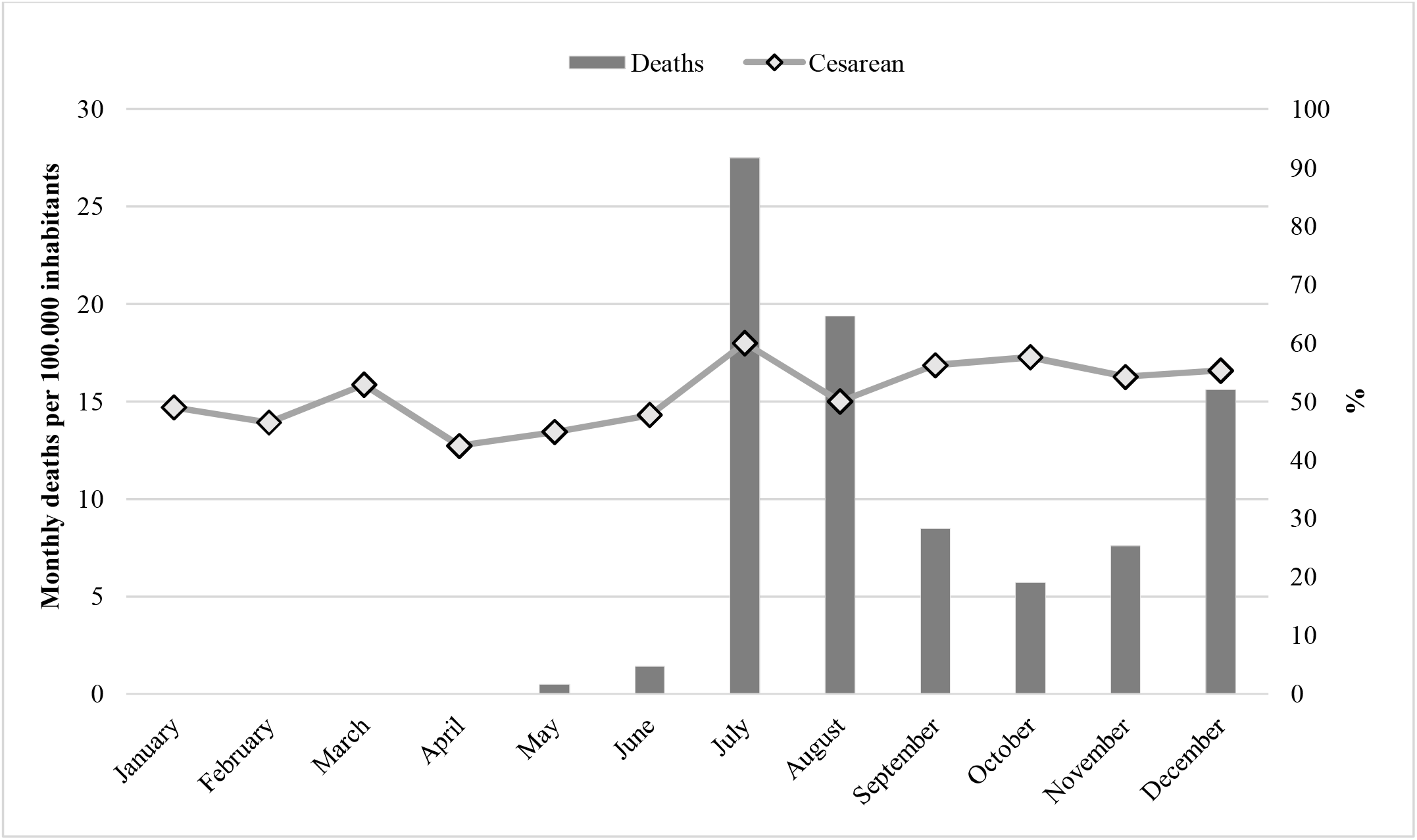
Prevalence of cesarean section in a baby-friendly hospital according to the number of deaths due to COVID-19 in the city of Rio Grande, RS, Brazil (2017-2020).

When comparing the cesarean prevalence ratios from 2017 to 2020, the data showed that in 2018/2017 there was no statistically significant difference in the prevalence ratio (95% CI: 0.95-1.14). In 2019/2018, there was a reduction of approximately 20% in the cesarean prevalence ratio (95% CI: 0.82-1.00). The scenario changes in 2020, with a 40% increase in cesarean prevalence compared to 2019 (95% CI: 1.16-1.42) (Table 3).

**Table 3.** Crude and adjusted prevalence ratios of cesarean section in a baby-friendly hospital. Rio Grande, RS, Brazil (2017-2020).

| Cesarean<br>/month | Prevalence Ratio (PR) |  |  |  |  |  |
| --- | --- | --- | --- | --- | --- | --- |
|  | 2018/2017 |  | 2019/2018 |  | 2020/2019 |  |
|  | PR crude<br>(CI 95%) | PR adjusted<br>(IC 95%) | PR crude<br>(CI 95%) | PR adjusted<br>(IC 95%) | PR crude<br>(CI 95%) | PR adjusted<br>(CI 95%) |
| January | 0.90 (0.63-1.28) | 0.85 (0.59-1.22) | 1.08 (0.76-1.54) | 1.05 (0.73-1.51) | 1.23 (0.87-1.74) | 1.23 (0.87-1.74) |
| February | 1.01 (0.69-1.47) | 1.05 (0.71-1.54) | 1.13 (0.77-1.67) | 1.10 (0.74-1.63) | 1.25 (0.86-1.82) | 1.25 (0.86-1.83) |
| March | 1.08 (0.78-1.51) | 1.10 (0.79-1.55) | 0.83 (0.59-1.18) | 0.82 (0.57-1.17) | **1.46 (1.04-2.04)** | **1.49 (1.06-2.10)** |
| April | 1.03 (0.75-1.40) | 1.02 (0.73-1.41) | 0.73 (0.51-1.04) | 0.75 (0.52-1.08) | 1.18 (0.82-1.70) | 1.14 (0.80-1.65) |
| May | 0.88 (0.64-1.20) | 0.86 (0.62-1.19) | 0.84 (0.60-1.19) | 0.87 (0.61-1.23) | 1.22 (0.85-1.76) | 1.22 (0.84-1.77) |
| June | 1.24 (0.90-1.71) | 1.17 (0.84-1.63) | 0.73 (0.52-1.03) | 0.77 (0.54-1.09) | 1.26 (0.88-1.80) | 1.27 (0.89-1.81) |
| July | 1.26 (0.94-1.70) | 1.26 (0.92-1.72) | 0.80 (0.59-1.09) | 0.82 (0.60-1.13) | **1.41 (1.01-1.95)** | 1.34 (0.97-1.85) |
| August | 0.95 (0.70-1.31) | 0.94 (0.68-1.30) | 0.97 (0.70-1.36) | 0.98 (0.69-1.38) | 1.19 (0.85-1.67) | 1.25 (0.89-1.77) |
| September | 0.88 (0.63-1.22) | 0.86 (0.62-1.20) | 0.86 (0.60-1.23) | 0.81 (0.56-1.16) | **1.55 (1.08-2.22)** | **1.59 (1.11-2.29)** |
| October | 1.15 (0.80-1.65) | 1.11 (0.77-1.60) | 0.92 (0.67-1.30) | 0.91 (0.65-1.29) | 1.33 (0.96-1.85) | **1.50 (1.07-2.10)** |
| November | 1.08 (0.77-1.53) | 1.09 (0.76-1.55) | 1.22 (0.85-1.74) | 1.22 (0.85-1.75) | 1.01 (0.71-1.44) | 1.01 (0.71-1.45) |
| December | 1.11 (0.80-1.54) | 1.10 (0.78-1.54) | 0.95 (0.67-1.35) | 0.97 (0.68-1.40) | 1.30 (0.90-1.86) | 1.25 (0.86-1.82) |
| Total (Jan-Dec) | 1.04 (0.95-1.14) | 1.03 (0.93-1.13) | 0.91 (0.82-1.00) | 0.91 (0.82-1.00) | **1.27 (1.15-1.40)** | **1.28 (1.16-1.42)** |

CI: confidence interval; Analyzes adjusted for maternal age, birthweight, and gestational age.

Figure 2 presents in graph format the data shown in the table 3. Only the 2020/2019 prevalence ratios were above the unit, demonstrating that 2020 was a year in which there was greater risk of cesarean section. Moreover, when comparing month by month, the prevalence ratios for March, July, and September 2020 were statistically significant in relation to the same period the previous year. Such differences were not observed for the same months when comparing 2019 with 2018 and 2018 with 2017.

**Figure 2.**
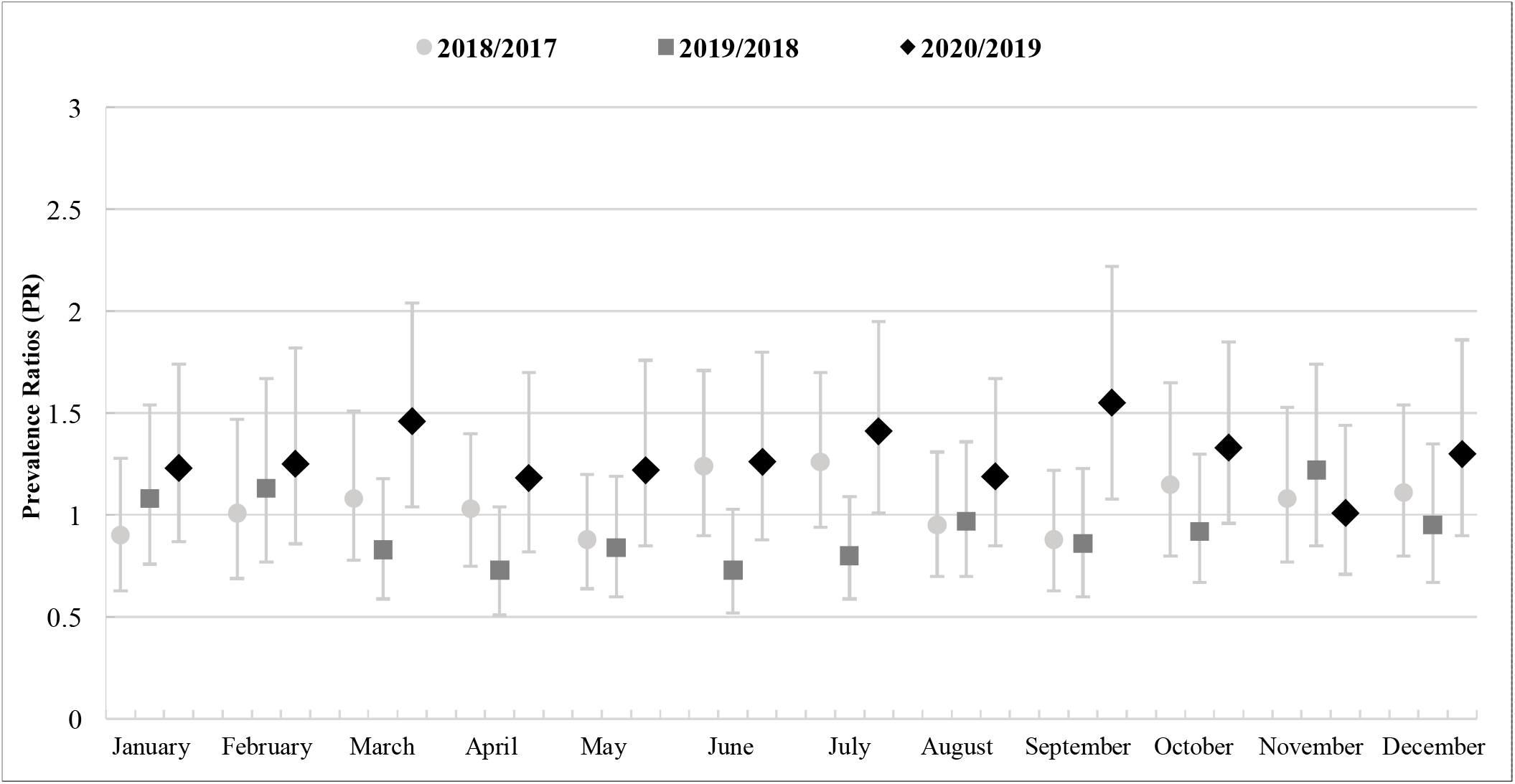
Cesarean section prevalence ratios in a baby-friendly hospital. Rio Grande, RS, Brazil (2017-2020).

## Discussion

The findings of this study reveal a significant increase in cesarean prevalence in 2020 in relation to the three preceding years. Especially when making a comparison with the year prior to the pandemic, cesarean prevalence raised from 40.2% in 2019 to 51.0% in 2020. This increase was pronounced in July 2020, in line with the first COVID-19 mortality peak in the municipality.

The increase in cesarean prevalence observed in the present study may have occurred because cesarian section can be considered a faster procedure for reducing the time pregnant women spend waiting for childbirth, and thus reducing the amount of time they are exposed to the hospital environment. Elective cesarean sections can be scheduled for a specific data and this reduces exposure between hospital admission and child delivery, whereas vaginal delivery and emergency cesarean sections need a team of professionals and hospital beds to be available. It is possible that the increase in cesarean sections can be related to deliveries that could have been vaginal delivery, whether this was the mother’s option or the medical team’s option. Notwithstanding, cesarean sections are related to longer postpartum hospital stay time, differently to vaginal delivery for which recovery is quicker.

It is important to note that the evolution of COVID-19 in the State of Rio Grande do Sul was different to that in other regions of Brazil, starting later in relation to other states and peaking in July. In the case of the municipality of Rio Grande, the first case of COVID-19 mortality was notified in May 2020^18^. Moreover, as HU-FURG is a public institution, it became the reference hospital for COVID-19 treatment in the municipality. Moreover, it is noteworthy that up to December 31st 2020 no cases of COVID-19 in pregnant women had been recorded on the hospital’s information system.

The World Health Organization (WHO)^19^ has summarized COVID-19 transmission routes, indicating that newborns can be contaminated by the virus during and after birth, either by direct transmission through contact with or breathing in respiratory secretions of infected people (family members and visitors), or though indirect transmission through contact with contaminated surfaces and objects^19,20^. Studies report uncertainties about contamination through vertical transmission of the virus from an infected mother to her baby, i.e. either during pregnancy through transmission of germ cells or placental blood, or at birth during delivery^21-25^. However, according to the United States Centers for Disease Control and Prevention, there is no evidence of SARS-CoV-2 being transmitted in this way^20,26^. In view of this, the care protocols for pregnant and puerperal women recommend considering normal obstetric indications and individual assessment of the severity of the mother’s condition when choosing the form of delivery, so that COVID-19 infection should not be considered as a reason for indicating cesarean section^15^. Nevertheless, studies have shown an increase in cesarean prevalence globally^22,27-29^.

The increase in cesarean prevalence during the pandemic observed in the present study is consistent with the findings reported in other countries^27,29^. In England, Bhatia et al. (2020) assessed six English hospitals where there was a total of 2,480 births between April and July 2020. The authors found a 29.7% increase in cesarean prevalence when compared to the same period in 2019 (28.3%)^27^. However, this trend in cesarean prevalence was not reported by Malhotra et al. (2020)^28^ in 11 New York hospitals where 1,952 births were recorded between March 8^th^ and April 20^th^ 2020. In that study, 31.3% of pregnant women who tested positive for COVID-19 were submitted to cesarean sections, and the prevalence rate was near the overall cesarean prevalence in preceding years which was around 31%. The authors also observed that cesarean prevalence in women who had not been tested for the virus was 31.4%, and among those who had tested negative it was 33.9%. Thus, these findings did not suggest that being contaminated with COVID-19 interfered in the decision as to which form of child delivery.

A recent study by Mor et al. (2021) assessed the impact of the pandemic on emergency obstetric care and on perinatal outcomes in a low-risk population in Israel. Authors identified that during the first wave of COVID-19 in Israel (from February to April 2020), emergency child delivery hospitalizations were lower on average when compared to the same period in the preceding years (2017-2019)^30^. Moreover, they found a significantly higher rate (p=0.037) of stillbirths during the study period. On the other hand, cesarean prevalence was not impacted in relation to the prevalence rates found in the previous years, which ranged from an average of 17.5% between 2017 and 2019 to 17.7% in 2020^30^.

It is also important to discuss the cesarean rates observed for years prior to the pandemic. Despite being high, the rates are similar to studies conducted in BFHI hospitals^31^. The “*Nascer no Brasil*” study assessed 22,035 puerperal women and their newborn babies and found 40.6% cesarean rates in BFHI hospitals compared to 58.9% in hospitals not accredited by the BFHI^29^. In the birth cohort studies conducted in Pelotas, a municipality neighboring Rio Grande, the overall cesarean prevalence observed in 2015 was 65.1%, being more than double compared to births in 1982 (27.6%)^32^. Thus, cesarean prevalence rates both in Brazil and in the extreme south of the State of Rio Grande do Sul are high, even in BFHI hospitals, and thus more actions will be needed to reduce the rise of surgical deliveries in the post-pandemic period.

This study has strong points that should be highlighted. The data analysis proposed used a large sample as well as data from four years to assess evolution of the results. As it is a secondary data study, the possibility of measurement bias is reduced, given that the electronic medical records and forms are filled in by trained hospital staff almost immediately after child delivery. Another aspect is the absence of selection bias, considering that the sample included all deliveries performed at the hospital, only excluding deliveries meeting pre-established exclusion criteria for indicating cesarean section.

Some limitations need to be considered. The first refers to the absence of information on the electronic system about the indication of cesarean section, whether due to medical indication or other factors, such as medical convenience or patient’s choice. Another limitation is external validity. The data presented refer to a municipality in the extreme south of Brazil and are not representative for cities and regions where the repercussions of COVID-19 have been alarming.

The results of this study show high cesarean prevalence rates in a BFHI hospital during the first year of the COVID-19 pandemic, worsening even more the cesarean section epidemic in Brazil. Mode of delivery and cesarean section are subjects widely discussed in the literature, as child delivery is a moment of extreme importance for mothers and their families, whereby each woman’s individual context must be considered. Repercussions of the pandemic on the mode of delivery are still scarce in Brazil. Further studies are needed to investigate possible changes in childbirth protocols in maternity hospitals in the context of the COVID-19 pandemic, especially in baby-friendly hospitals. We emphasis the need to plan actions to keep cesarean prevalence within BFHI recommended rates to minimize the deleterious effect of the pandemic in possible excessive cesarean section and future consequences for maternal and child health.

## Data Availability

The dataset supporting the conclusions of this article is not openly available due to confidentiality of information. Consent for publication of raw data was not obtained but a fully anonymous dataset can be obtained with the corresponding author.

